# Long-term Mortality Among Hospitalized Adults with Sepsis in Uganda: a Prospective Cohort Study

**DOI:** 10.1101/2023.09.14.23295526

**Authors:** Paul W. Blair, Stephen Okello, Abdullah Wailagala, Rodgers R. Ayebare, David F. Olebo, Mubaraka Kayiira, Stacy M. Kemigisha, Willy Kayondo, Melissa Gregory, Jeff W. Koehler, Randal J. Schoepp, Helen Badu, CDR Nehkonti Adams, Prossy Naluyima, Charmagne Beckett, Peter Waitt, Mohammed Lamorde, Hannah Kibuuka, Danielle V. Clark

## Abstract

**Background:** Twelve-month mortality in sepsis survivors has not been previously characterized in sub-Saharan Africa.

**Methods:** Hospitalized adults with ≥ 2 modified systemic inflammatory response syndrome (SIRS) criteria (temperature < 36°C or > 38°C, heart rate ≥ 90 beats per minute, or respiratory rate ≥ 20 breaths per minute) were enrolled at a tertiary care centre from October 2017 to August 2022. Multiple clinical blood and respiratory molecular and antigen assays were used to identify infectious etiologies. Baseline demographics were evaluated for risk of death by 1 month and 12 months using Cox proportional hazards regression.

**Results:** Among 435 participants, the median age was 45.0 years (interquartile range [IQR]: 28.0, 60.0) years, 57.6% were female, and 31.7% were living with HIV. Malaria (17.7%) followed by tuberculosis (4.7%), and bacteremia (4.6%) were the most common detected causes of illness. Overall, 49 (11.3%) participants died, and 24 participants died between one month and one year (49.0% of deaths and 5.5% of the cohort). Female participants had a decreased risk of death by 12-months (unadjusted hazard ratio [HR]: 0.37; 95% confidence interval [CI]: 0.21 to 0.66).

**Conclusions:** The burden of sepsis may be underestimated in sub-Saharan Africa due to limited long-term follow-up.

## Introduction

In 2017, 11 million deaths occurred due to sepsis^1^. The burden of sepsis is highest in low- and middle-income countries (LMIC), settings where treatment resources are lowest ^1^. Long-term mortality after sepsis is high, ranging from 28 to 66% over 12 months ^2^. However, long-term outcomes after sepsis in LMICs are largely unknown with only one, to our knowledge, prospective study past 28-days ^3^ and the overwhelming majority of studies coming from North American and Europe. There have been no published studies from sub-Saharan Africa (SSA) describing post-sepsis mortality at 12-months ^2–5^.

We conducted a prospective observational study to characterize severe infectious disease pathogens, clinical manifestations, and host response among patients presenting for care at Fort Port Regional Referral Hospital in Fort Portal, Uganda^6^. One purpose of the study has been to build a continuous base of severe infectious disease clinical research operations and training in preparation for more complex protocols, including interventional clinical trials^6^. We describe long-term (12-month) outcomes after five years of enrollment, causes of sepsis, and the mortality risk associated with baseline clinical characteristics.

## Materials and Methods

Persons 18 years of age and older presenting with suspected infection and ≥ 2 modified systemic inflammatory response syndrome (SIRS) criteria (temperature < 36°C or > 38°C, heart rate ≥ 90 beats per minute, or respiratory rate ≥ 20 breaths per minute) were enrolled at Fort Portal Regional Referral Hospital (FPRRH) in Fort Portal, Uganda. SIRS was incorporated in sepsis cohorts^7^ prior to Sepsis 3 criteria ^8^ and maintained for consistency. Adult patients 18 years of age and older presenting at the outpatient department, emergency department, or medical wards were evaluated for enrollment from October 2017 to August 2022. Patients were not eligible if they were deemed too ill to participate with an imminently terminal comorbidity or if they presented with severe anaemia (hemoglobin<7 g/dL) due to phlebotomy requirements. Due to initial inflammation biomarker objectives^9^, participants were excluded with known immunocompromising conditions including drug induced immunosuppression, anatomic or functional asplenia, recent chemotherapy, pregnancy and<6 weeks post partum females, but participants with HIV were eligible. FPRRH serves as a health facility to eight districts in western Uganda (Supplementary Figure S1) and includes an isolation unit for referral of patients with high consequence pathogens including during the 2022 Sudan virus disease outbreak ^6,10,11^. After screening potential participants, 435 participants enrolled from October 2017 to August 2022. The study team collected demographic, clinical, and laboratory data at 0, 6, and 24 hour time points after enrollment and at 72 hours starting after the first 260 enrollments. Study participants were followed until discharge and at 1 month when a physical exam, history, and labs were also done. Telephone follow-ups were done at 6-months and 12-months. All clinical and laboratory data were recorded daily by designated study staff on the case report forms. Survival was determined during in-person visits or telephone calls when participants could not attend in-person visits. There were 398 participants at 28-days follow-up, 383 participants at 6-month follow-up, and 338 participants at 12-month follow-up (Figure 1).

**Figure 1.**
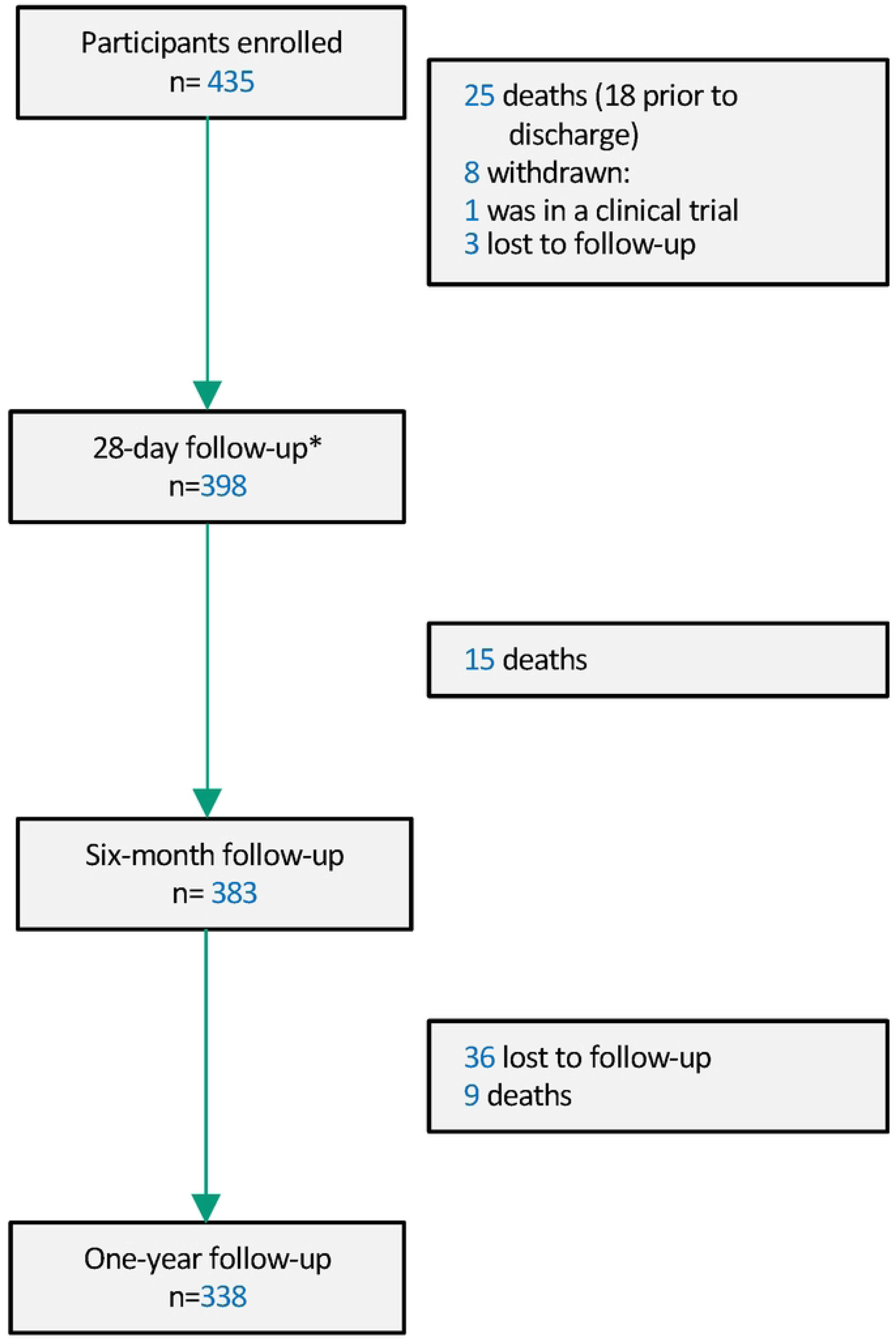
Cohort flow diagram.

Physiologic parameters (heart rate, blood pressure, oxygen saturation, and temperature) were prospectively collected at study visits at enrollment, 6 hours, and 24 hours while hospitalized and at in-person follow-up visits. After enrollment, an i-STAT analysis (Abbott, Chicago, IL) for lactate and blood gases was performed on blood collected at scheduled at 0, 6, and 24 hour study visits. Clinical tests were routinely performed including complete blood counts, coagulation analyses, and chemistries. Diagnostic testing included blood culture with antimicrobial sensitivity testing, urinalysis, HIV testing with consent, malaria smears and malaria rapid diagnostic tests were routinely performed. If there was blood culture growth, the BCID FilmArray (Biofire, Salt Lake City, Utah, USA) assay was run for bacterial species confirmation. If there was clinical suspicion based on respiratory symptoms, testing for tuberculosis was performed using PCR sputum testing (Xpert MTB/RIF Ultra, Cepheid, California, USA). Patients with known COVID-19 were not enrolled until June 2021. Hospital-based COVID-19 testing (rapid diagnostic tests or PCR) was recorded until replaced by multiplex PCR sputum testing (Xpert® Xpress SARS-CoV-2/Flu/RSV, Cepheid, California, USA) in July 2022 for those with respiratory symptoms. If HIV testing was positive, urine lipoarabinomannan (Abbott TB LAM Ag, Illinois, United States) testing was performed. The Biofire FilmArray Global Fever polymerase chain reaction (PCR) panel (Biofire, Utah, USA) was run on whole blood samples to identify 19 bacterial, viral, or protozoal targets or plasma if whole blood was unavailable (first 36 participants) ^12^. Plasma samples were run in duplicate with a Crimean-Congo hemorrhagic fever virus real-time RT-PCR assay targeting the NP gene using previously described methods ^13–15^ but replaced after transition of the Global Fever panel from off-site to on-site use after the first 237 participants. Extracted nucleic acid (5 μl) from plasma samples from the first 237 participants were run with a Crimean-Congo hemorrhagic fever virus real-time RT-PCR assay targeting the NP gene in duplicate using the using SuperScript One-Step RT-PCR Kit (ThermoFisher Scientific) using previously described methods ^13–15^. Primers (CCHFV-S-F649 5’GGAVTGGTGVAGGGARTTTG and CCHFV-S-R705 5’-CADGGTGGRTTGAARGC) were at 5 μM concentrations, and the probe (CCHFV-S-p670 FAM-CAARGGCAARTACATMAT-MGBNFQ) was at 0.1 μM M. Cycling conditions were 50°C for 15 min; 95°C for 5 min; and 45 cycles at 94 °C for 1 s, 55 °C for 20 s, and 68 °C for 5 s. Fluorescence was measured after each extension step, and a sample was considered negative if the quantification cycle (Cq) was greater than 40 cycles.

### Analysis

Summary statistics were performed for baseline characteristics and microbiology results. Baseline demographics were compared between those that completed followed to those that did not (were withdrawn or lost-to-follow-up, 6.4%) using the Kruskal-Wallis test. Person-time was administratively censored at the known last visit among those who were lost-to-follow-up or withdrawn. After checking the proportional hazards assumption using Schoenfeld residuals and log-log plots (resulting in exclusion of platelet count and dichotomizing respiratory rate), baseline characteristics of age, sex, HIV status, clinical laboratory results, qSOFA score were evaluated for their association with 28-days and 12-month survival with Cox regression models. The logrank test was performed to evaluate for a difference in survival based on HIV status or sex. Sepsis severity clinical decision tools were dichotomized for bivariate models according to current usage, including qSOFA score ≥2 (range, 0 [best] to 3 [worst] points)cand UVA score ≥2 (range, 0 [best] to 13 [worst])^16^. Bivariate analyses were exploratory; therefore, a p-value correction was not performed.

Enrollment in the Fort Portal sepsis cohort to a sample size up to 1,075 participants is planned to estimate the risk of emerging pathogens with a precision of 3%. We herein present results from the first 435 participants to disseminate clinical epidemiologic information, inform clinicians about the differential diagnosis of severe infection in Uganda, and describe long-term outcomes after the first five years of enrollment. A map of Uganda was created using ArcGIS ArcGIS Online (Redlands, CA)^17^. Analyses were performed using Stata, version 16.0 (StataCorp LLC, College Station, TX, USA), and figures were created using Stata or R, version 4.0.1 (R Foundation).

## Results

Participants were a median (IQR) of 42.0 (28.0 to 57.0) years of age at enrollment and 59.1% were female (n= 257) (Table 1). A diagnosis of human immunodeficiency virus (HIV) was common (31.7%; n=138) of which 17.4% (n=24) were new diagnoses. Otherwise, known comorbid illness was uncommon (Table 1). At enrollment, there were 21.2% (n=91) with a qSOFA score of ≥2. Tachypnea and tachycardia was common but most participants were normotensive and afebrile (Table 1). The median duration of hospitalization was 3.0 days (IQR: 2.0 to 5.0 days).

### Diagnoses, clinical microbiology, and antibiotics

Among the 32.4% (141 of 435 participants) with a positive microbiologic result, malaria was the most common cause of illness. There were 6.3% (27 of 424) with positive malaria smears and 17.7% (75 of 425) with positive malaria rapid diagnostic tests (Table 3). Among 43 RDT positive samples with smear quantification results, the median parasitemia was 10,240 (IQR: 1,680, 60,000) parasites/μL of blood. The Global Fever PCR panel was most frequently positive from *Plasmodium spp.* (25.4%; 102 of 401) or specifically *P.falciparum* (19.5%, 78 of 401).

Non-malarial positive Global Fever panel PCR results were uncommon but included two cases of Leptospira *spp.*, and one case of *Salmonella enterica* serovar *Typhi*. A single positive CCHF virus result was obtained with single-plex RT-PCR and confirmed with national referral laboratory testing at the Uganda Viral Research Institute. Intrerestingly, this CCHF case was a 25-year-old woman with one week of fevers and chills with no hemorrhagic symptoms, a normal complete blood count, and only a borderline elevated AST (30 U/L) on the complete metabolic panel. Hospital-based COVID-19 testing was positive in 32.7% (35 of 107 participants). Results including a Delta variant surge^18^ from June 2021 to December 2021 were positive in 58.3% (28 of 48 participants tested) but positivity decreased to 11.9% (7 of 59 participants tested) from January 2022 to August 2022.

The most frequent confirmed non-malarial causes of sepsis were tuberculosis and bacteremia. Of 160 participants able to produce sputum samples, 10.6% (17 of 146 with sputum) were positive for *M. tuberculosis* by Cepheid GeneXpert. Among those with HIV and urine samples, 5.0% (5/121) had positive LAM results including 3 without sputum Cepheid GeneXpert testing resulting in 20 total tuberculosis cases (4.6% of the cohort). Out of 414 participants with blood cultures, there was growth among 4.6% (N=19) of participants and *Streptococcus pneumoniae* (N=7; 1.7%) was the most common isolate (Table 2). One out of 1 cerebrospinal fluid cultures grew *Cryptococcus neoformans.* The most common antimicrobials received among participants included ceftriaxone (85%), metronidazole (34.7%), azithromycin (27.8%), artesunate (17.5%), and ciprofloxacin (17.2%). Notably, 7.8% received anti-tuberculosis treatment and 6.4% received doxycycline or tetracycline. Among those with microbiologic testing, 32.9% (141 of 428 participants) had a positive result. Majority (36 of 49, 73.5%) of deaths did not have a microbiologic diagnosis. Among those with positive results, testing indicated tuberculosis (n=3), malaria (n=3), bacteremia (n=4) and COVID-19 (n=4).

### Risk of death at 28-days and 12-months

Overall, 49 (11.3%) participants died. By 28-days, 25 (5.7%) participants died, and 24 deaths occurred between 28-days and 12-months (49.0% of deaths and 5.5% of the cohort). Among those with a qSOFA ≥2 at enrollment, 18.8% (17 of 91 participants) died by 12-months. Compared to the rest of the cohort, those lost-to-follow-up had a similar age (median 44, IQR: 33.5 to 59.0 years; p=0.29), female sex (50.0%; p=0.31), prevalence of HIV (25.0%, p=0.43) and baseline qSOFA score (median 1, IQR: 1 to 1; p=0.90). Death occurred a median of 46.0 (IQR: 7.0 to 178.0) days from enrollment and 38.8% of those that died had HIV. In bivariate analyses, while age was not associated with increased risk of death, female participants had a lower risk of death over 28-days (HR: 0.44;95% CI: 0.21 to 0.75; Supplementary Figure S2A) and 12-months (HR: 0.37; 95% CI: 0.21 to 0.66, logrank test p<0.001, Figure 2A and Figure 3A) than male participants. People with HIV were not observed to have an increased risk of death over 28-days (HR: 1.02; 95% CI: 0.44 to 2.36) or 12-months (HR: 1.38; 95% CI: 0.77 to 2.44; logrank test p=0.19). Baseline physiologic metrics were associated with an increased risk of death over both 28-days and 12-months including an increased heart rate, decreased oxygen saturation, decreased mental status, qSOFA score ≥2, and UVA score ≥2 (Figure 2C-D, Figure 3A). Clinical laboratory parameters associated with increased risk of death over 28-days and 12-months included decreased serum sodium, increased neutrophil/lymphocyte ratio, increased AST, increased creatinine, and increased lactate (Supplementary Figure S2B and Figure 3B). A positive malaria RDT was not associated with a difference in risk of death over 28-days (HR: 0.40; 95% CI: 0.09 to 1.70) but was associated with a decreased risk of 12-month mortality (HR: 0.30; 95% CI: 0.09 to 0.96)(Figure 3C). Any microbiologic diagnosis was not associated with a difference in risk of death over 28-days (HR: 0.96, 95% CI: 0.41 to 2.23) or 12-months (HR: 0.73, 95% CI: 0.39 to 1.37).

**Figure 2.**
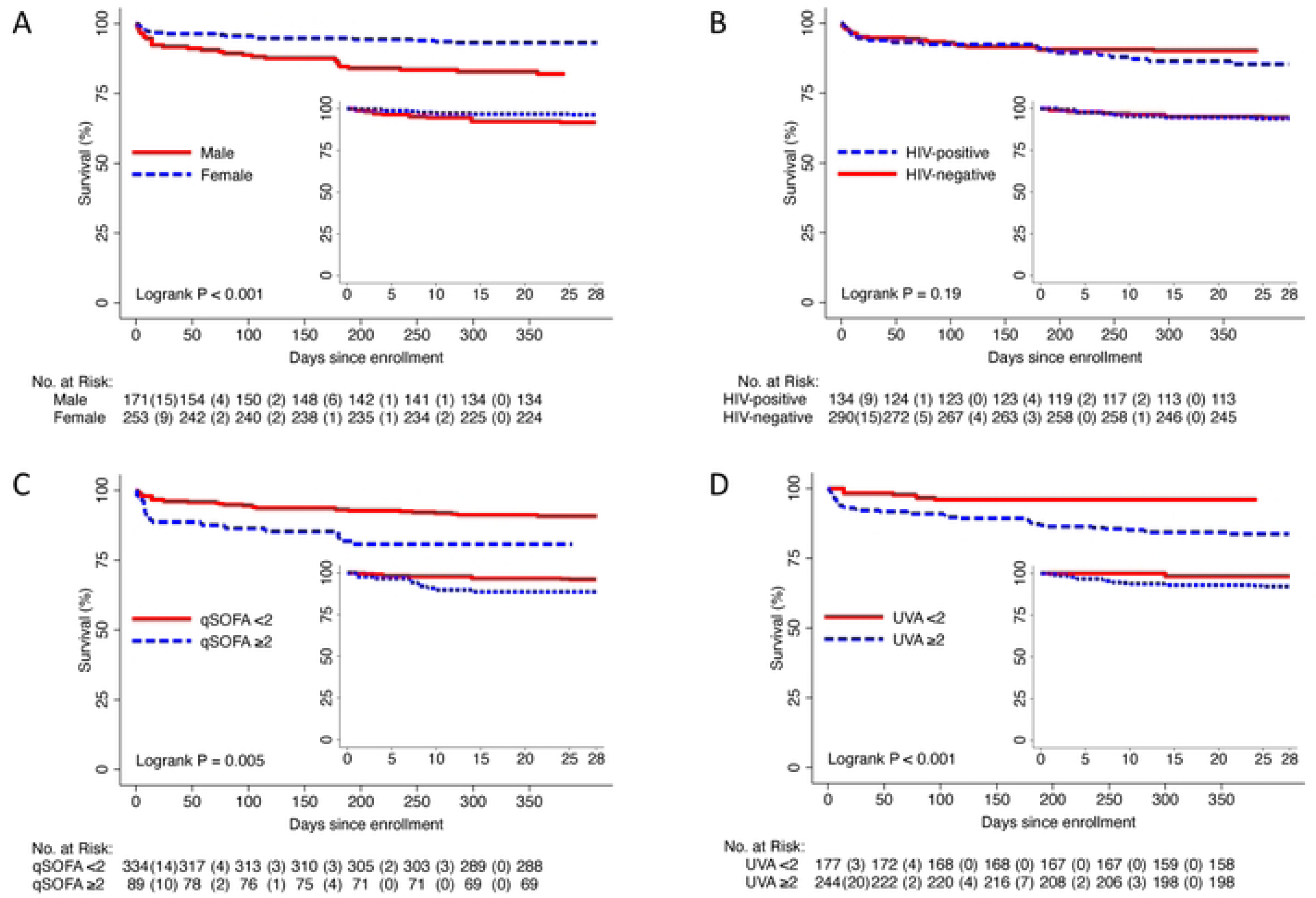
Kaplan-Meier plots by sex (A), HIV status (B), qSOFA score (C), and UVA score (D).

**Figure 3.**
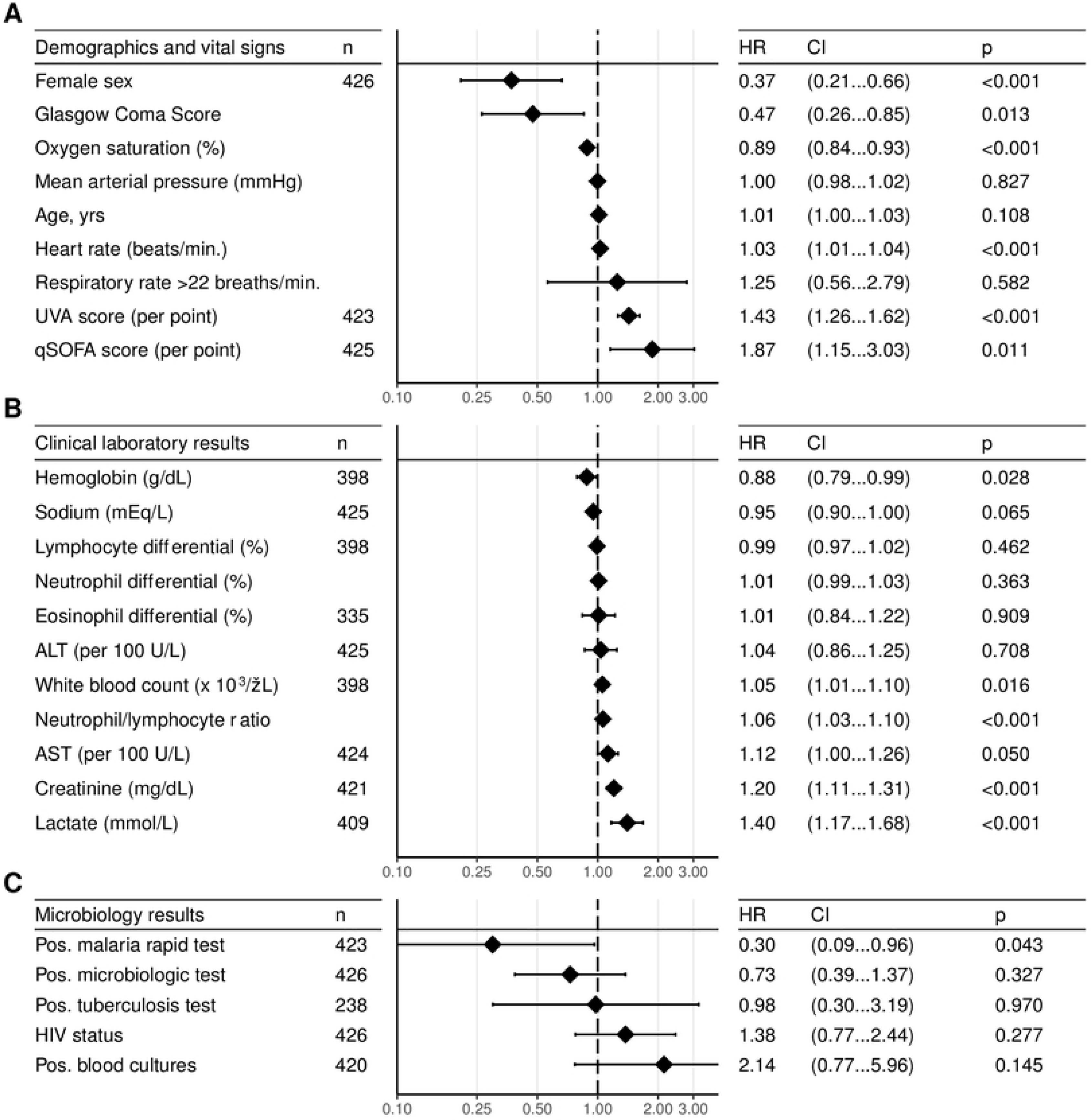
Hazard ratio forest plot of 12-month mortality risk associated with baseline demographics and vital signs (A), clinical laboratory results (B), and clinical microbiology results (C). Min.:Minute, qSOFA: quick Sepsis Organ related Failure Assessment; Pos.: positive; UVA: Universal Vital Assessment.

## Discussion

Our study is the first to describe mortality at one year after generalized sepsis in a well-characterized large prospective cohort in sub-Saharan Africa (SSA) ^3,5^. We identified that almost half of deaths among adults that presented with suspected sepsis occurred after 28-days. The most common causes of illness were malaria, tuberculosis, and bacteremia, and an unsuspected case of CCHF was identified. COVID-19 was the most common cause of illness during a 2021 surge. The majority of causes of illness were unknown despite multiple clinical microbiologic assays.

There is a paucity of long-term outcome data in SSA. In a meta-analysis of 15 studies of sepsis in SSA performed in 2019, none had results of mortality after 30 days ^5^. Compared to the 11.3% 12-month mortality we observed, the pooled in-hospital sepsis mortality among those meeting SIRS score of ≥2 from 7 cohort studies with enollment years ranging from 2008 to 2013 was 19% (95% CI, 12 to 29%, n=1159) ^5^. HIV status was not associated with a difference in risk of mortality, consistent with a recent small acute febrile illness cohort in Uganda with a similar baseline prevalence of an elevated qSOFA score ^19^. The similar risk of death observed among those with HIV is potentially a testament to outcome improvements from increased access to outpatient antiretroviral treatment ^19,20^. However, this may not be generalizable to other countries in SSA. Patients with HIV had a higher risk of death in a recent sepsis cohort in Malawi^3^. In that study, death was observed in 18% at 28-days and in 31% at 180 days. Overall lower death rates in our cohort may have been due to baseline patient population differences, including nonspecificity of SIRS criteria for sepsis eligibility^7^, or advancements in clinical care at the Fort Portal site ^6^. Long-term mortality may be higher in more severely ill individuals or in other settings with less resources for sepsis management. Standardized multi-site cohorts and surveillance studies are needed for definitive conclusions.

Although sepsis syndrome is likely a final common pathway for many of these infectious diseases-related deaths, few studies from SSA have provided insight into the microbiologic diagnoses of these patients. A meta-analysis of African bacteremia studies reported a 10% prevalence of bloodstream infections ^21^. A more recent 2019 meta-analysis demonstrated that tuberculosis dominates as a cause of sepsis in SSA ^5^. Similar to our findings, standard clinical microbiological laboratory analysis for bacterial, mycobacterial, and malarial pathogens were detected a cause in only ∼30% of cases, most commonly *Mycobacterium tuberculosis*, non-typhoidal Salmonella, *Staphylococcus aureus*, and *Streptococcus pneumoniae*. A recent study of adult and paediatric patients hospitalized with acute febrile illness in Tanzania were discovered to have acute bacterial zoonoses as the most common group of aetiologies (26.2%) including leptospirosis, Q fever, and spotted fever group rickettsioses, none of which are identified by standard culture methods ^22^.

Our findings highlighted the public health utility of ongoing infectious illness cohorts. Our study identified a case of CCHF that otherwise would not have been detected. Similarly, CCHF was previously identified at the Global Fever panel febrile illness study site in Mubende, Uganda ^12,23^. During the 2022 Sudan virus disease outbreak. multiple suspect Ebola cases have been identified as CCHF. Detection of these cases suggests that CCHFV is an uncommon but persistent cause of suspected sepsis or febrile illness in Uganda and that cases without fulminant hemorrhagic symptoms likely occur undetected outside of outbreaks. Additionally, we identified COVID-19 as the most common causes of suspected sepsis in this remote referral hospital during a local surge^18^. Our finding emphasize the unaccounted burden of admissions during outbreaks and subsequent deaths due to outbreaks and emerging infectious diseases in low-resource and remote settings. As emerging infectious disease threats become more commonplace and with challenges from known endemic threats, highlighted by the 2022 Sudan virus outbreak in Uganda ^11^, our need to rapidly recognize and effectively manage these causes of severe infection grows more urgent. In recent years, several public health emergencies of international concern were first detected in critically ill patients, patients meeting the diagnostic criteria for sepsis. Scaling and developing point-of-care diagnostics would help facilitate early recognition of emerging causes of sepsis to initiate infection control, supportive treatment, and reporting of emerging infectious diseases.

We identified several clinical parameters associated with death by 28-days and by 12-months. Consistent with prior studies ^3,19^, decreased oxygen saturation was found to be associated with an increased risk of death. Patients with respiratory failure may be at particularly risk in hospitals without respiratory support including high flow nasal cannula and mechanical ventilation or access to supplemental oxygen upon discharge. Elevated UVA scores, qSOFA scores, and point-of-care lactate were each associated with an increased risk of death consistent with prior studies in LMICs ^7,16,24^. Prior research that demonstrated the additive prognostic value of serum lactate in Uganda, Malawi, and Southeast Asia ^3,25,26^. A positive malaria result was associated with a decreased risk of death at 12-months. While this finding is tempered by multiple comparisons, a protective association was observed in the Malawi cohort ^3^. Malaria in adults with suspected sepsis may be less fatal due to longstanding immunity, access to diagnostics, and commonly employed empiric anti-malarial treatment regimens. Detection of any pathogen was not associated with risk of death but inference is complicated by intrinsic pathogen differences, effects on treatment decisions, and increased assay sensitivity with higher inoculum infections. Future research should be performed to evaluate the effect of point-of-care diagnostics for decreasing time to accurate treatments.

Our study has several limitations. First, while laboratory testing in this cohort was robust and included a broad molecular panel, blood cultures, and rapid diagnostic tests, tuberculosis and rickettsial infections were likely missed but current point-of-care diagnostics are limited for these pathogens. CD4 or HIV viral load results were not routinely available, preventing inference about level of HIV control. Second, the effect of sepsis on quality of life at one month was not ascertained at follow-up visits. Future research plans to include surveys to determine the long-term burden of sepsis on quality of life. Third, despite similar baseline characteristics, those lost to follow-up may have been more at risk for death; our estimate of long-term mortality may be underestimated. Despite limitations, to our knowledge this is the first cohort study in SSA to describe long-term mortality to 12-months after suspected sepsis.

## Conclusions

Almost half of the deaths observed overall occurred during the observation period from one month to one year. The burden of sepsis in SSA is likely underestimated. Patients discharged after admission for sepsis likely represent a vulnerable population that should have a longitudinal follow-up for medical care.

## Declarations

This protocol and informed consent were approved by the U.S. Army Medical Research and Development Command Institutional Review (# M-10573) and Makerere University School of Public Health IRB# 490). All participants provided written consent that was provided in either English or their local language. All procedures were in accordance with the ethical standards of the Helsinki Declaration of the World Medical Association. The investigators have adhered to the policies for protection of human subjects as prescribed in 45 CFR 46.

The contents of this article are the sole responsibility of the authors and do not necessarily reflect the views, assertions, opinions, or policies of the Henry M. Jackson Foundation for the Advancement of Military Medicine, Inc., the U.S. Department of Defense, the U.S. government, or any other government or agency. Mention of trade names, commercial products, or organizations does not imply endorsement by the U.S. government. Some of the authors of this work are military service members or employees of the U.S. government. This work was prepared as part of their official duties. Title 17 U.S.C. x105 provides that ‘‘Copyright protection under this title is not available for any work of the United States government.’’ Title 17 U.S.C. x101 defines a U.S. government work as a work prepared by a military service member or employee of the U.S. government as part of that person’s official duties.

## Data Availability

De-identified data will be made available upon reasonable request and if it is in accordance with local IRB regulations.

## Acknowledgments

We would like to thank all of the Fort Portal research nurses, laboratory technicicians, and hygienists and the medical staff. We would also like to thank David Brett-Major, MD, James Lawler, MD, Nahid Bhadelia, MD, and Karen Martins, PhD for their contributions to the Fort Portal research site.

## Author contributions

PWB conceived and designed the study, analyzed, and interpreted data, and wrote the manuscript. DFO, AW, SO, and MK collected data. SO, MG, RJS, PW and HB contributed to project execution. ML, CB, HK, and DVK oversaw project execution and contributed to obtaining resources. PN, AW, DFO, HK, RJS, and PW contributed to interpretation of results. All authors reviewed and approved this manuscript.

